# Automating the analysis of eye movement for different neurodegenerative disorders

**DOI:** 10.1101/2023.05.30.23290745

**Authors:** Deming Li, Ankur A. Butala, Laureano Moro-Velazquez, Trevor Meyer, Esther S. Oh, Chelsey Motley, Jesús Villalba, Najim Dehak

## Abstract

The clinical observation and assessment of extra-ocular movements is common practice in assessing neurological disorders but remains observer-dependent and subjective. In the present study, we propose an algorithm that can automatically identify saccades, fixation, smooth pursuit, and blinks using a non-invasive eye-tracker and, subsequently, elicit response-to-stimuli-derived interpretable features that objectively and quantitatively assess patient behaviors. The cohort analysis encompasses persons with mild cognitive impairment (MCI) and Alzheimer’s disease (AD), Parkinson’s disease (PD), Parkinson’s disease mimics (PDM), and controls (CTRL). Overall, results suggested that the AD/MCI and PD groups exhibited significantly different saccade and pursuit characteristics compared to CTRL when the target moved faster or covered a larger visual angle during smooth pursuit. When reading a text passage silently, more fixations were an AD/MCI-specific feature. During visual exploration, people with PD demonstrated a more variable saccade duration than other groups. In the prosaccade task, the PD group showed a significantly smaller average hypometria gain and accuracy, with the most statistical significance and highest AUROC scores of features studied. The minimum saccade gain was a PD-specific feature distinguishing PD from CTRL and PDM. Furthermore, the PD and AD/MCI groups displayed more omitted antisaccades and longer average antisaccade latency than CTRL. These features, as oculographic biomarkers, can be potentially leveraged in distinguishing different types of NDs in their early stages, yielding more objective and precise protocols to monitor disease progression.

## I. Introduction

Neurodegenerative disorders (NDs) encompass a number of progressive neurological disorders that manifest with central nervous system degeneration and present with impairments of movement, coordination, mood, and/or cognition. Advancing age strongly correlates with the risk of developing NDs, representing a tremendous socioeconomic and personal burden, particularly with lifespan increases in many countries [1]. Common NDs include Alzheimer’s disease (AD), Parkinson’s disease (PD), Frontotemporal Dementia, and a number of other related or mimicking conditions. AD is, both, the most common ND and the most common form of dementia worldwide, followed by Vascular Dementia and Dementia with Lewy Bodies [2], while PD is the most common neurodegenerative movement disorder that affects 2–3% of the population ≥65 years of age worldwide [3], [4].

Many NDs have a prodromal stage. Persons presenting with mild cognitive impairment (MCI) may, in some cases, be in prodromal stages of AD, before more severe memory and linguistic decline occur. The diagnosis of MCI due to AD is usually given to people with MCI who have biomarker positivity for AD (e.g., abnormal brain positron emission tomography (PET) scans or cerebrospinal spinal fluid biomarkers). In contrast, PD is an ND defined on primarily clinical terms based on the presence of bradykinesia combined with either rest tremor, rigidity, or both. Notably, the clinical presentation of both AD and PD is multifaceted and includes many overlapping nonmotor symptoms [5]. Lastly, co-pathology is common [6], [7]. However, the gold standard test for a definitive diagnosis of PD or AD requires pathology evidence of neurodegeneration via autopsy. Guided by the awareness of disease subtypes, treatment plans vary from person to person, emphasizing the need for precision medicine and personalized management.

The complex and multifaceted presentation of NDs means that accurate diagnosis may require months or years. A major goal of current clinical research in NDs is improving early and accurate disease detection, which would facilitate implementing treatment as early as possible and assess disease progression [8]. Many approaches are being undertaken to identify biomarkers to improve early detection, which opens the gate for accelerated and more accurate neurological diagnosis in conjunction with other biomarkers like imaging, biomedical and genomic profiling. Specifically, algorithms have been developed to provide gait analysis to monitor symptoms in PD [9]. Acoustics in terms of articulatory and phonatory aspects of speech and voice are used to support automatic detection and severity assessment of PD [10]. More speech-based acoustic, linguistic, and cognitive biomarkers were proposed to discriminate between subjects with NDs, including PD, AD, and other PD-related disorders [11], [12]. Handwriting is another biosignal that could reveal fine motor deficits in different NDs based on kinematic, fluency, and micrographia analysis [13].

In this work, we use non-invasive eye tracking to record eye motion and gaze location across time and tasks. Eye tracking data contains not only rich information on eye movement but also provides quantitative insights into brain functioning. In a review of oculomotor features of the major age-related movement disorders, Anderson and MacAskill [14] suggested that oculomotor signs can be leveraged for differential diagnosis in NDs, such as PD [15], [16], [17], AD [18], [19], spinocerebellar ataxia [20], [21], [22], and Huntington disease, [24], [25]. The clinical assessment of extraocular muscle (EOM) function includes, at minimum, an examination of the ability of individuals with NDs to fixate, track a moving target, and perform saccadic eye movements. More complex physiological tasks, such as antisaccades, memory-guided saccades, and repetitive to-and-fro saccades, among others, are often included on a case-by-case basis [26]. In previous studies, AD patients were found to have delayed initiation of saccade for fixation, along with shorter fixation periods than healthy controls [27], [28]. Pavisic et al. suggested that, during smooth pursuit tasks, the AD group spent less time pursuing the target compared to age-matched healthy controls [28]. Saccadic eye movement abnormalities in AD were observed when patients had higher latency and latency variability for saccade initiation, less accurate prosaccade, and a larger number of saccades to fixate the target [29], [28], [30]. Moreover, Noiret et al. [29] showed that AD patients made more uncorrected antisaccade and had a longer latency to initiate a corrective saccade in the antisaccade task.

Antoniades and Kennard [31] suggested that the most consistent ocular motor abnormality in PD is saccadic hypometria, in which the primary saccade undershoots the target, especially vertically [32], [33], [34]. There are also deficits in the initiation and performance of self-regulated tasks, including antisaccades and memory-guided saccades [33], [35], [36], [37]. Voluntary saccade execution dysfunctions and difficulties inhibiting reflexive saccades in PD were reported in the work by Amador et al. [38], such that PD patients showed more errors in the antisaccade task and an increased number of disinhibitions in the delayed antisaccade task. These results are consistent with an impairment of the frontal-basal ganglia circuits that leads to deficits in the control of voluntary saccade generation [39]. Furthermore, repeated trials of mixed-up pro and antisaccade tasks in a block increased prosaccade and antisaccade error rates for PD patients compared to single-task blocks [40]. Participants with PD produced more saccades during smooth pursuit than the control group, and some of them also showed impaired binocular coordination, reported in [41]. Tsitsi et al. [42] observed that the median pupil size and the longest fixation period differed between PD and healthy controls. Moreover, De Boer et al. [30] found that AD, PD, and control subjects exhibited significantly different visuomotor behaviors in eye-hand-coordination.

One limitation of the available literature is the assessment of a singular ND using a single or a few extraocular tasks. By analogy, this would be akin to attempting to diagnose a condition based on a single or few examination findings. In contrast, the strength of the current study is that we analyze the behavior of a group of features in different NDs (AD/MCI, PD, and Parkinson’s Disease Mimics [PDM]) and a control group employing multiple distinct tasks. Second, most of these studies include 20 or 30 minutes of recording on a single task instead of our protocol which favors faster, more efficient methods. In our case, five tasks (36 trials in total) were used, which took under 15 minutes to complete in a multi-modal multi-test rapid battery. Third, a meta-analytic review of 38 articles in the literature [17] reports a mean of 14.2 participants per study for PD and 11.8 for controls. The total population (n=143), especially the number of controls (n=58) and the number of PD (n=41), included in this study is relatively large in the topic of eye-tracking metrics for evaluating NDs, bolstering the significance of our findings. Lastly, manual extraction of eye-tracking features or biomarkers is time-consuming, which makes this practice unfeasible to analyze a patient’s condition in clinical practice. Hence, it becomes advantageous to automatically analyze eye movement to identify eye motion states and extract interpretable quantitative features, shedding light on the functioning of different parts of cognitive systems. Our analysis is implemented in an automated pipeline that can accomplish analysis with minimal human supervision ^1^. The method is intended to assist clinicians in assessing the presence and monitoring the disease progression of different NDs. Fig. 1 illustrates a diagram of our automated pipeline.

**Fig. 1.**
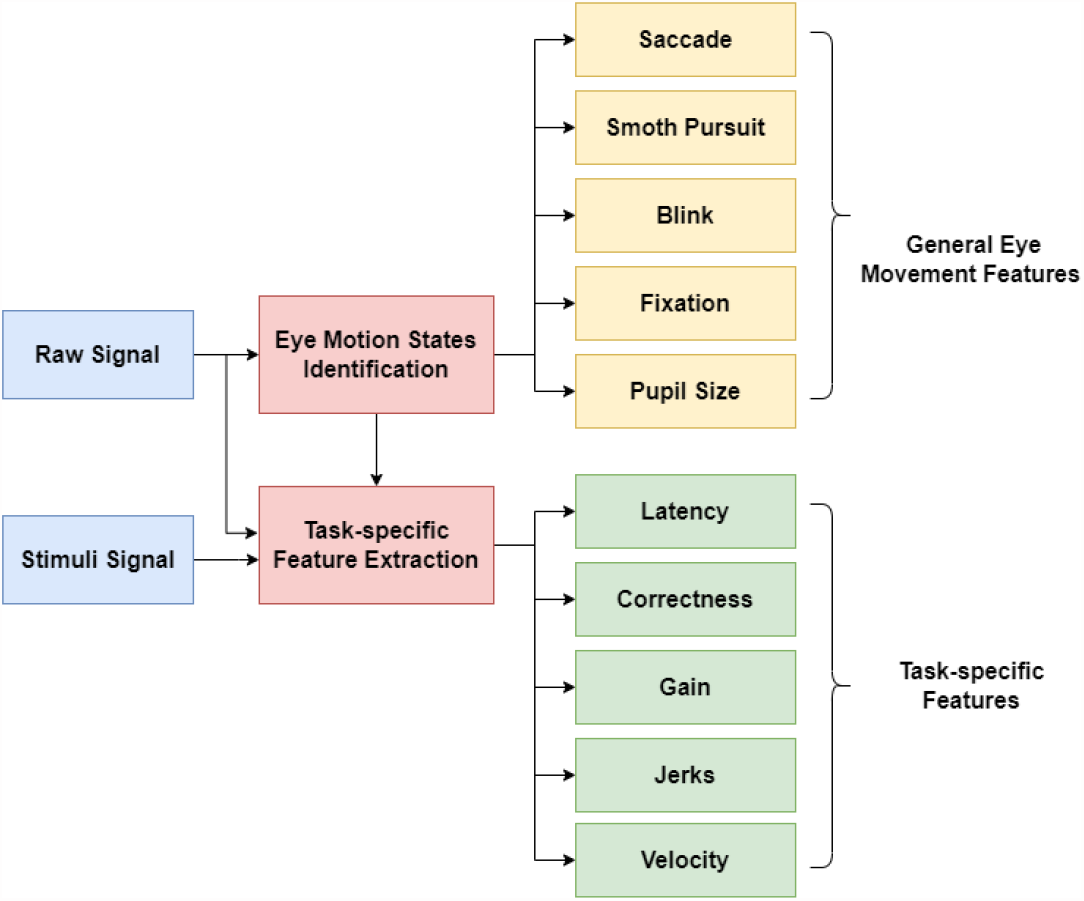
A block diagram of the main modules in our automated pipeline

## II. Materials

### A. Data set

*NeuroLogical Signals* is an ongoing multi-modal corpus being collected by the authors of this study and currently contains 143 participants, including control subjects (CTRL), people with AD or MCI, PD, and various PDM. All ND patients were seen at Johns Hopkins Medicine, and all participants signed informed consent. The data collection was approved by the Johns Hopkins Medical Institutional Review Board. AD and PD were selected since these are the two most prevalent neurodegenerative diseases. Comparing these two groups will allow us to observe if any of the proposed features (potential biomarkers) are AD- or PD-specific. The AD [43], PD [44], and PDM [45], [46], [47] groups contained patients with diagnoses that were *clinically established*. Fourteen participants with MCI, of which six were biomarker-positive for AD, were combined with six AD patients forming the AD/MCI group to create a larger group. All participants in the PD group had a predominant clinical syndrome of Parkinsonism. The PDM group encompassed varying degrees of PD-related movement disorders, atypical Parkinsonism, and secondary Parkinsonism, including Progressive Supranuclear Palsy [45], Dementia with Lewy Bodies [47], Corticobasal Syndrome [48], and Multiple System Atrophy [46]. In this study, all PDM participants were initially determined to have physical findings of parkinsonism and met, at some point in their work-up, “Possible PD,” but whose diagnosis evolved over subsequent visits. While inclusive of multiple etiologies, the PDM group contains persons who could be misdiagnosed with Probable PD, and it acts as an active control group for the PD cohort. Whereas our goal is not to find any specific knowledge about this heterogeneous group, finding features that could differentiate between PD and PDM is highly relevant [49]. The table I shows the cohort study’s demographics and disease severity statistics. We report sample size, sex, age distribution, and scores on the Montreal Cognitive Assessment (MoCA) for each experimental group. In addition, we report clinical dementia rating scale sum of boxes (CDR-SB) for the AD/MCI group and unified Parkinson’s Disease Rating Scale Part III (MDS-UPDRS III) for the PD and PDM groups.

**TABLE I.** NLS Data Set.

| Data set | Category | Sample (n) |  |  | Age |  | MoCA |  | CDR-SB |  | MDS-UPDRS III |  |
| --- | --- | --- | --- | --- | --- | --- | --- | --- | --- | --- | --- | --- |
|  |  | tot | female | male | avg | range | avg | range | avg | range | avg | range |
| NLS | CTRL | 58 | 29 | 29 | 67.60 (8.83) | 38-83 | 25.9 | 16-30 | - | - | - | - |
|  | AD/MCI | 20 | 5 | 15 | 70.84 (8.2) | 57-84 | 19.6 | 6-28 | 4.0 | 0-11 | - | - |
|  | PD | 41 | 13 | 28 | 66.7 (10.9) | 40-86 | 25.5 | 21-30 | - | - | 24.3 | 8-44 |
|  | PDM | 24 | 12 | 12 | 62.25 (8.2) | 49-81 | 24.7 | 18-30 | - | - | 39.3 | 14-74 |

### B. Data collection

The eye tracking data was recorded with EyeLink Portable Duo by SR Research. The setup included an eye-tracking unit with a camera and an infrared torch, a host PC connected to the camera and dedicated to data acquisition, and a display PC on which stimuli were presented during experiments. The display screen was 380×215 mm (1920×1080 in pixels). The distance from the participant’s head to the screen was 500 mm, on average. We employed the head-free mode in which a sticker was positioned in the middle of the participant’s forehead to localize their eyes. This mode was preferred over head-fixed since the latter requires chin rests, which was problematic for some participants with postural and motoric problems. Moreover, a chin rest would interfere with the tasks that require the participant to speak, as described in Subsection II-C. We asked the participants to try to remain as stable as possible while completing the visual tasks. The EyeLink system provided the coordinates of where the eyes looked on the screen (gaze) based on the detected pupil and corneal reflex, with a sampling rate of 1000 Hz per eye. In all tasks, the image background was grey (R=G=B=153/255), and the targets were of high contrast. The screen luminance was maintained, whereas the environmental light was very similar across all the participants. Calibration and validation were performed at the beginning of the session, and drift correction was performed before each task to ensure the system was calibrated. The average validation error for each group was within one visual degree. The raw signal provided by the system was composed of time sequences of [*x, y*] positions, velocities, and pupil relative size. The stimulus signal could also be extracted from eye-tracking data files to enable task-specific analysis.

### C. Tasks

Five tasks, illustrated in Figure 2, were performed to assess the ocular motor and cognitive behavior in NDs, i.e., Smooth Pursuit, Prosaccade, Antisaccade, Cookie Thief (visual exploration), and Rainbow Passage (read text). We hypothesized that these tasks could produce different features indicative of motor and cognitive-related factors, which can be correlated clinically. Every task was analyzed with the eye motion states identification algorithm to find the saccades, smooth pursuits, blinks, and fixations within the recording. Using that raw data and identified eye motion states, we extracted a set of general eye movement features and a set of task-specific features. The tasks were intended to be at an appropriate difficulty level, avoiding saturatingly large number of errors from being too difficult and limited effects of measures from being too easy [50]. The tasks listed were selected to provide a complementary screen of cognitive dysfunction common to NDs: Smooth Pursuit (motion perception), Prosaccade (vigilance, alertness, motoric aspects of the saccade generation system), Antisaccades (sustained attention, error detection, task sustainment), Cookie Thief (visual attention/monitoring v. inattention/neglect, spontaneous language generation, attentional shifts), and Rainbow Passage (reading, dictation, articulation, phonation, etc.).

1. *Smooth Pursuit:* The participants followed a target red dot with a diameter of 22 pixels, equivalent to 0.5° visual angle across the screen. Seven smooth pursuit trials with different stimulus moving patterns were performed (horizontal line: n = 2; vertical line: n = 2; infinity pattern: n = 3). The target movement pattern is sinusoidal, to avoid abrupt changes in direction and speed. The oscillation frequencies were 0.2 and 0.4 Hz, horizontally and vertically in the first four trials. The amplitude of the two horizontal trials (1 and 2) was 14°, and the amplitude of the two first vertical trials (3 and 4) was 8°. The target frequency amplitude of the infinity patterns varied in the last three trials. The detailed configurations are summarized in the Appendix, Table III. Each trial lasted 18 s.
2. *Prosaccade:* Participants were initially presented with a green dot with a diameter of 0.5° visual angle in the center of the screen. After this, two target red dots of the same size were subsequently shown in opposite directions, either left or right horizontally or up or down vertically. The participants were asked to look at the target as quickly and accurately as possible when it appeared without moving their heads. The targets appeared in possible locations: ±2°, ±4°, ±6°, ±9°, ±10°, ±11°, or ±18° randomly interleaved and presented equal or unequal distance in opposite directions. The prosaccades task comprised eight horizontal and eight vertical trials, which were mixed in order. The target locations, stimuli time, and orders were different for each trial but constant for all participants. This means that the time a green or a red dot was on the screen varied between 900 and 2100 ms depending on the trial to avoid temporal predictability of target onset [50], but this time and the order of the trials were the same for all participants.
3. *Antisaccade:* Similar to the Prosaccade task, the participants were initially presented with a green point subtending at 0.5° visual angle in the middle of the screen. After this, a yellow point was shown either to the right or left side of the screen horizontally or to the top or bottom of the screen vertically. The participants were asked to look at the opposite region of the screen. The possible target locations were ±9°, ±10°, ±14°, ±16°, or ±18° in the horizontal direction and ±4°, ±5°, ±8°, or ±9° in the vertical direction. The yellow point lasted variously, ranging from 900 ms to 2100 ms. The resting time between each trial also varied between 900 ms to 2100 ms. These settings were constant across all participants. Ten horizontal trials followed by ten vertical trials were performed in this task. The main purpose of the antisaccade task is to test disinhibition, which is defined as “the inability to withhold a prepotent response or suppress an inappropriate or unwanted behavior. It can refer to the production of socially inappropriate comments and/or actions.” [51]
4. *Cookie Thief:* As a task of self-directed visual exploration, the participants were asked to describe the description of a picture within one minute [52]. The picture depicts a familiar scene from everyday life with distinct characters, activities, and place contrasts [53]. No prompting was provided regarding what to describe, such that they were encouraged to talk about anything and everything in the scenery. Elementary keywords were expected to be used when describing the scene. Cookie Thief has been one of the most used tasks to elicit disclosure abilities and is particularly useful in assessing the integration of cognitive–linguistic abilities [53], [54]. The results of the speech analysis of this and other similar tasks in our cohorts can be found in [11], [12].
5. *Rainbow Passage:* The participants read the Rainbow passage [55], with two variations: 1. the participants read the first half of the passage out loud. 2. The participants read the rest of the passage to themselves instead. The rainbow passage is short and phonetically balanced. It contains a variety of sounds element and is commonly used as a standardized test to assess a person’s speech abilities, such as pronunciation and enunciation. This work analyzes eye movement to extract patterns and provides insights into how the participants perform while completing this task.

**Fig. 2.**
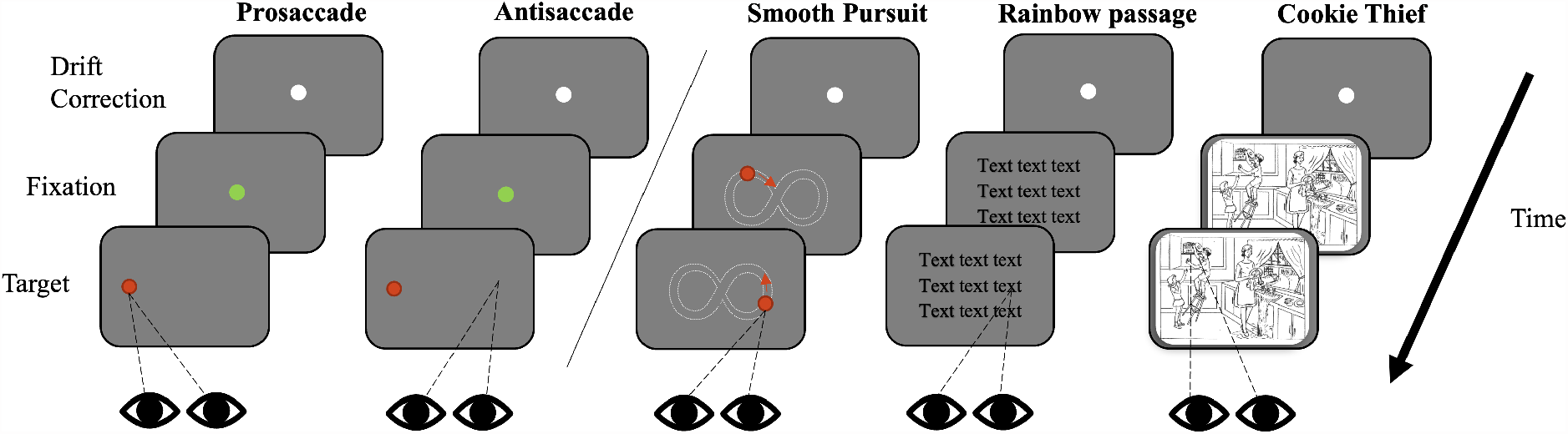
Example sequences of screens for the five tasks employed along the timeline. Prosaccade and Antisaccade are almost identical, except the participant is instructed to saccade towards or away from the target. Smooth Pursuit involves visual tracking of a target with a predetermined pattern (infinity sign in this example). Cookie Thief involves the visual exploration of an image with a domestic scene.

## III. Methods

We developed an algorithm based on distance, velocity, and acceleration thresholds to determine eye motion states from eye-tracking data automatically. Then, two groups of eye movement interpretable features were extracted to evaluate the motoric and cognitive patterns in the cohorts. Finally, we studied the statistical significance of the differences between groups (AD/MCI, PD, PDM, and CTRL) employing the Kruskal-Wallis H test [56] and Benjamini–Hochberg correction [57].

### A. Eye Motion States Identification Algorithm

Four eye motion states, i.e., saccade, smooth pursuit, blink, and fixation, are typically considered when analyzing eye movement. A saccade involves a rapid eye movement from one point to another, often lasting within 100 ms [58], and the velocity is commonly above 30°/s [59]. Smooth pursuit involves tracking stimulus as it moves with a slow and steady eye movement. A blink involves a rapid closing and opening of the eyelid. Fixations are eye movements that stabilize the retina over a stationary object of interest [60], often lasting more than 100 ms. Because we cannot expect smooth pursuit if there are no moving targets on the screen, smooth pursuit movement is only analyzed with our algorithm in the smooth pursuit task.

The data contained time series of gaze position on a screen [*x, y*] (pixels), velocity [*v*_*x*_, *v*_*y*_] (°/s), and pupil size for two eyes. The eye with the lowest validation error was selected for analysis. Horizontal and vertical accelerations [*a*_*x*_, *a*_*y*_] were computed from [*v*_*x*_, *v*_*y*_] respectively based on the equations:

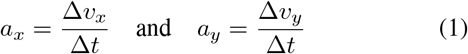

where Δ*v* is the change in velocity, and Δ*t* is the change in time. The overall velocity and acceleration were calculated by:

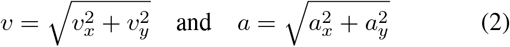

The goal of our algorithm 1 is to identify the eye motion states by determining the corresponding starting and ending points in the eye-tracking data.

- Saccade segments: To determine in which region of the data there is a saccade, we used thresholds of 40 pixels (0.91°) for saccade distance (TSD), 30°/s for saccade velocity (TSV), and 6000°/*s*^2^ for saccade acceleration (TSA). Initial saccade candidate segments were determined by finding the regions where velocity was above TSV and merged if gaps between two consecutive candidate saccade segments were shorter than 20 ms. The start and end points were extended for each candidate segment while the acceleration was larger than TSA. Similar approaches have been discussed in the Eyelink User Manual and [59]. After the extension, the candidate segment was identified as a saccade if the distance between the start and end was larger than TSD to discard microsaccades and artifacts.
- Smooth pursuit segments: To determine in which region of the data there is a smooth pursuit, we used 50 pixels thresholds (1.1°) for smooth pursuit distance (TPD) and a lower-bound 5°/s in the velocity (TPV), which was intended to filter out small changes in the gaze signal. A new saccade velocity threshold of 60°/s [61] (TSPV) was set to separate saccades from the background velocity of smooth pursuit eye movement. Therefore, a candidate segment was identified as smooth pursuit movement if velocity was consistently greater than TPV and lower than TSPV during the movement and the distance between the start and end was larger than TPD.
- Blink segments: To determine in which region of the data there is a blink, the candidate segments were evaluated by checking if no eye position was detected. Consecutive blinks with gaps less than 50 ms were merged. Because closing and opening of the eyelid cause disturbance in detecting the gaze position near the beginning and end of the blink due to the blocked pupil, the algorithm was configured to classify the blink segments from rise to drop in the gaze position so that the entire segment is recognized.
- Fixation segments: All segments that were not saccade, smooth pursuit, or blink were labeled fixations.

In each trial, the data was removed during quality checking if a summed duration of blinks or missing gaze coordinates was greater than 20 percent of the total trial length.

Fig. 3 shows several event graphs generated by the eye motion states identification algorithm for a participant and three tasks. Identified blinks were marked in yellow, saccades in red, and fixations in blue, whereas solid lines represent the gaze positions and dashed lines represent target positions. Fig. 3 (a) shows the gaze coordinates of one eye of a participant who followed a target point in motion describing the infinity pattern in the smooth pursuit task. In the prosaccade task (Fig. 3 (b)), the reaction time feature, marked with a black bar, indicates the delay to initiate a reflexive saccade after the stimulus appears. It includes an example of the participant with two hypometric saccades, each followed by one corrective saccade. The participant in Fig. 3 (c) performed a saccade in the correct, contralateral direction during the antisaccade task after an initial impulsive/erroneous prosaccade. Stepped saccades and occasional fixations are shown in (d) for the Rainbow Passage read task. In (e), we can see the identified saccades and fixations in image exploration for the Cookie Thief task. The event graphs of all the tasks and participants were visually checked to validate the output of the algorithm.

**Fig. 3.**
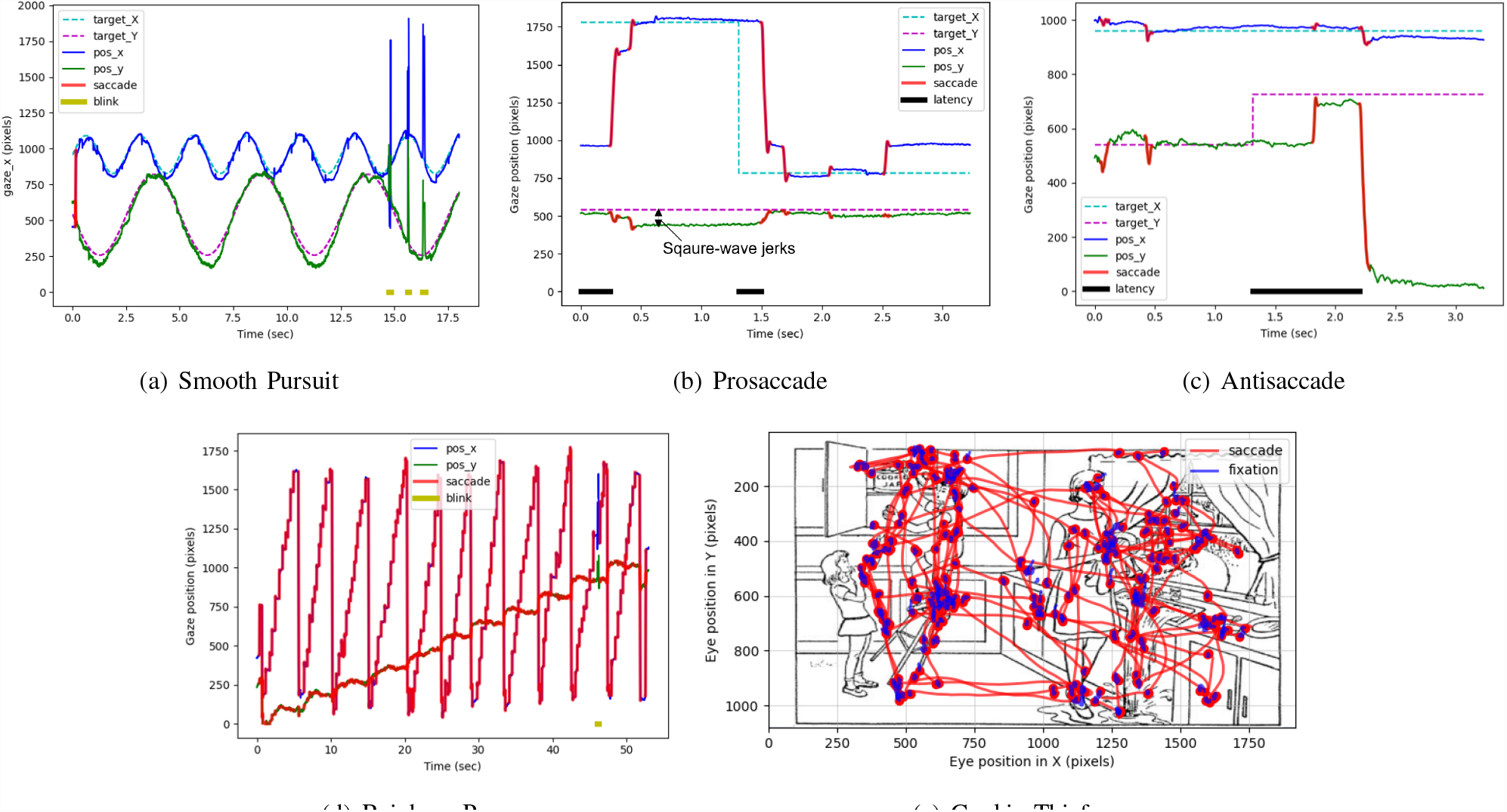
Visualization of identified eye motion states. (a), (b), (c), and (d) are plots of eye movement position vs. time. For a better view, the Cookie Thief task is visualized in the 2D gaze plane superimposed on the task image in (e).

### B. Feature Analysis

Two sets of interpretable features, listed in Table II, were designed. The first set was based on general eye movement related to saccades, smooth pursuit, blinks, fixations, and relative pupil size changes, without requiring the information of stimulus to compute. The other set of features depended on the interaction with stimuli, i.e., requiring stimulus position to calculate, specifically for smooth pursuit, pro, and antisaccade tasks. Following the internationally standardized prosaccade and antisaccade protocol by Antoniades et al., [50], we measured latency in saccade initiation, the gain of the prosaccade, hypometria or hypermetria percentage, errors (incorrect saccade or no response), and peak saccadic velocity. In addition, we measured the variability of eye movement by the number of corrective after the first saccade and the square-wave jerks. These sets of eye-tracking metrics could reveal the motoric patterns in different NDs as well as cognitive patterns, especially in the antisaccade task, which was cognitively more demanding to suppress the reflexive saccades.

**TABLE II.** Summary of the extracted general eye-movement and task-specific features. The description for each feature was reported. Abbreviations: PS, prosaccade, AS, antisaccade, #, number.

| Task | Features | Analysis type |
| --- | --- | --- |
| **General eye-movement** |  |  |
| All | Saccade count [#], duration [ms], distance [pixels], and velocity [$^{\circ}$/s] | Motoric |
| All | Smooth pursuit count [#], duration [ms], distance [pixels], and velocity [$^{\circ}$/s] | Motoric |
| All | Blink count [#], distance [pixels], and duration [ms] | Motoric |
| All | Fixation count [#] and duration [ms] | Motoric |
| All | Relative change in pupil size [%] | Motoric |
| **Task-specific** |  |  |
| Pursuit | Difference between the gaze and target positions [pixels] | Motoric |
| Pursuit | Pursuit gain [%] | Motoric |
| PS, AS | Latency of prosaccade or antisaccade [ms] | Motoric/Cognitive |
| PS, AS | Saccades in the wrong direction count [#] | Motoric/Cognitive |
| PS, AS | Errors of omission [#] | Motoric/Cognitive |
| PS, AS | Ratio [%] | Motoric/Cognitive |
| PS | Saccade gain, hypometria gain and percentage, hypermetria gain and percentage, and accuracy [%] | Motoric |
| PS | Corrective saccades [#] | Motoric |
| PS, AS | Square-wave jerks [pixels] | Motoric/Cognitive |

1. *General Eye Movement Features:* After saccades, smooth pursuits, blinks, and fixations were identified, the features of each eye motion state, including segment duration, distance, and velocity, were extracted. Then, for each trial and participant, all features were characterized by count, mean, maximum, median, and standard deviation statistics. For instance, the ideal case in a smooth pursuit trial would constitute only one complete segment of smooth pursuit movement from start to end, but it is also likely that a participant may have a certain count of smooth pursuit segments caused by blinks or intrusive and catch-up saccades. Similarly, in the Rainbow Passage task, a participant will have a certain number of fixations and saccades, which will also have a mean and max velocity. Because Pro and Antisaccade tasks had multiple similar trials based on the direction target was moving, i.e., horizontal or vertical, each was further split into two groups. As some previous studies indicate that subjects with PD have different horizontal and vertical saccade properties [32], [33], [34], we hypothesized that they might not perform consistently inter-trials based on direction. Lastly, since the pupil area information in our data had arbitrary units not calibrated across patients according to the Eyelink manual, we calculated and analyzed the relative percentage change in pupil size, which is unit-independent, between subsequent points instead of absolute values. The equation is as follows:

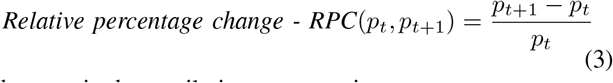

where *p*_*t*_ is the pupil size at every timestamp t.
2. *Task-specific Features:* The task-specific feature extraction algorithm was employed in the smooth pursuit, prosaccade, and antisaccade tasks. To quantify a participant’s response to stimuli, the features extracted from the smooth pursuit task were:
  1. Difference between the gaze and target coordinates. The smaller the difference is, the more accurate tracking of the stimulus.
  2. Pursuit gain is calculated by the ratio between the eye and the target velocity at every timestamp during the smooth pursuit eye movement, excluding saccades and blinks. Because the eye gaze velocity gradually reaches zero when approaching the turnaround points, to dismiss these parts of eye movement, only ratios greater than a cut-off threshold of 0.5 were considered pursuit gain [28].

In prosaccade and antisaccade tasks, developing upon the recommended outcome measures from Antoniades et al. [50], we measured a new set of features, specifically:

1. Latency of the correct prosaccade or antisaccade measures the response time from the target’s appearance to the saccade’s initiation. We hypothesized that NDs would demonstrate a greater latency to initiate a pro or antisaccade after stimuli presentation than controls.
2. Errors include the number of saccades in the wrong direction and the number of omissions (no response to the stimulus presented). For antisaccades, participants were supposed to look away from the stimulus. In this case, an incorrect saccade is made if the participant looks toward the stimulus. Also, an omission occurs if no saccade is detected relative to the stimulus. We hypothesized that the antisaccade task would be useful to differentiate the AD/MCI and the CTRL groups due to the added cognitive loading to inhibit a prepotent, i.e., prosaccade towards stimuli [50].
3. Gain of the prosaccade represents the ratio of actual saccade amplitude to target amplitude. We calculated the minimum, maximum, and standard deviation of the gain. We further defined that the prosaccade is a hypometria saccade if the prosaccade undershoots the target position by at least 1.5° or a hypermetria saccade if overshoot by at least 1.5° [28]. The average hypometria gain and hypermetria gain were derived based on this separation. We also calculated hypometria or hypermetria saccade frequency by the percentage of their respective count over the total prosaccade count; The accuracy of prosaccade was found by 1 − hypometria percentage − hypermetria percentage, which means that the attempted prosaccade falls within the 1.5° range of the target position. Since we did not ask the participants to make “mirror” saccades in the antisaccade task, i.e., to the exact opposite location on the screen, the gain of antisaccade, therefore, was not measured.
4. We further investigated the ratio of features by dividing them horizontally over vertically due to the possible different behaviors that may happen in different dominant directions.
5. Corrective saccades happens after a hypometria or hypermetria saccade. This feature counts the number of saccades after the initial saccade until the stimulus disappears.
6. Square-wave jerks of the gaze position in the non-dominant direction. Its magnitude is calculated by the distance between the eye gaze position and the target position, marked as the double arrow in Fig. 3 (b). For example, in the horizontal Pro or Antisaccade task, the vertical direction is the non-dominant direction, and vice versa. The jerks could be caused by inaccurate saccades and, ideally, should be as small as possible.
7. sPeak saccadic velocity and saccade duration of the pro and antisaccade.

## IV. Results and Discussion

We applied our eye motion state identification algorithm and extracted the features introduced in Section III. The results are summarized in Appendix, Table IV and explained in the subsections below. We also indicate the number of subjects per task in this table, as this number is not constant across tasks due to the fact that few participants, particularly in the AD/MCI group, recorded multiple sessions during revisits or noise during data collection. For each pairwise feature, we report the corresponding p-values based on *H-statistic* [56] to determine if there were any statistically significant differences between the experimental groups. The Kruskal-Wallis H test is a nonparametric test whose null hypothesis is that the mean ranks of the groups are the same. To control the false discovery rate (FDR) in many features, we applied Benjamini–Hochberg correction to each pair-wise comparison [57]. The error rate, *α*, was set to 0.05. We also report the area under the ROC curve (AUROC), which can be used as a criterion to measure the feature’s discriminative ability.

## A. General Eye Movement Features

Fig. 4 includes the distributions of the most relevant general eye movement features, i.e., those providing better separability between groups. We also define that a feature is ND-specific when its mean significantly differs from that of CTRL and at least one other ND group. We frame the distribution plots for those features in orange (PD-specific) or blue (AD-specific). For example, the standard deviation of saccade duration feature in Cookie Thief is a PD-specific feature as it is significantly different (*p <* 0.05) from other groups and unique to PD. When the active control group (PDM) is clearly different from the other groups, we mark the distribution in red.

**Fig. 4.**
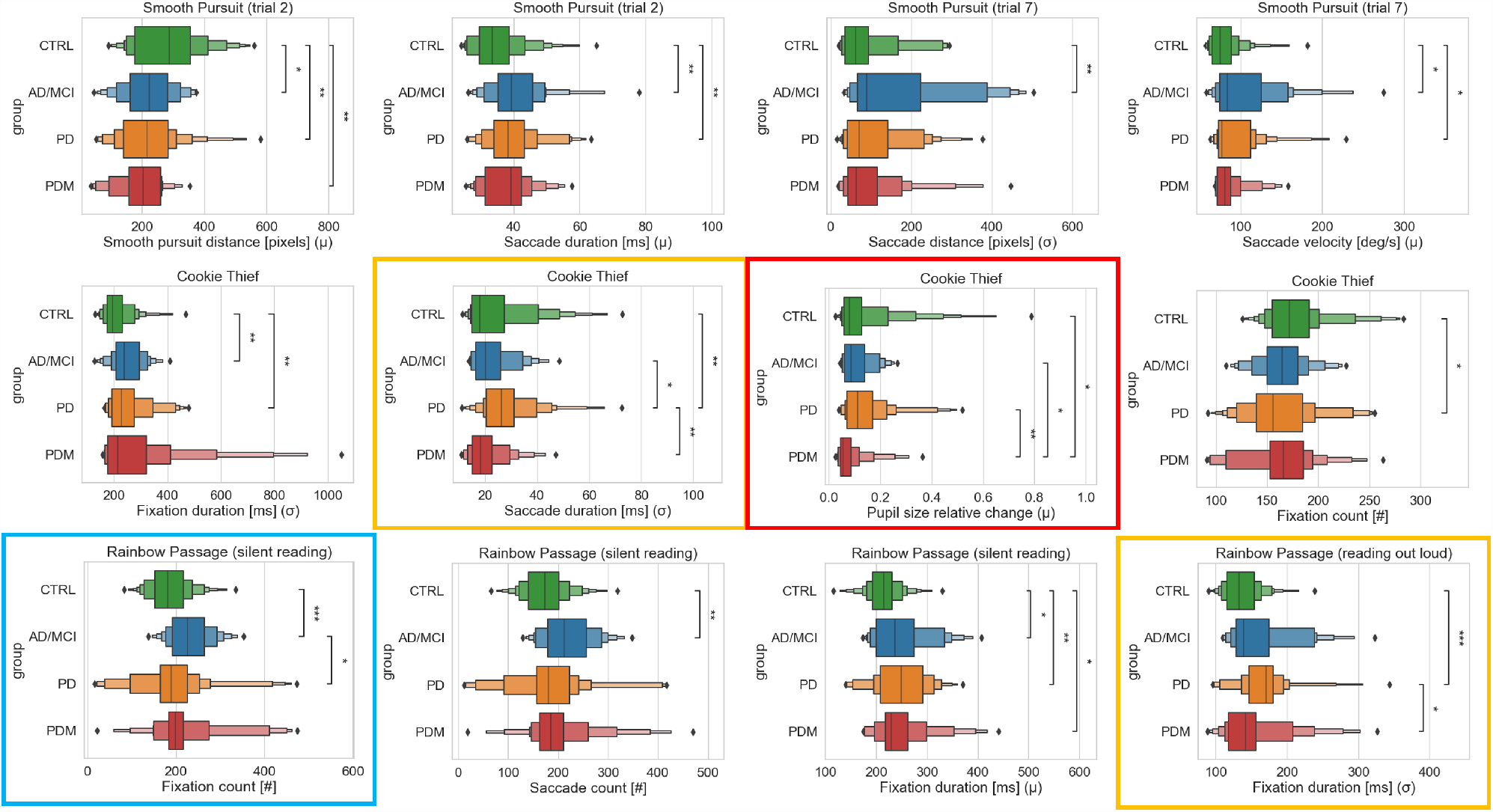
Comparison of general eye movement features between CTRL, AD/MCI, PD, and PDM using boxenplots (or letter-value plots [62]). The center line corresponds to the median. The data space is divided further recursively, and more quantiles are added to the depth while the corresponding letter values are reliable estimates. The outliers are represented with a diamond symbol. Asterisks indicate significant differences: ^*\**^*p <* 0.05, ^*\*\**^*p <* 0.01, ^*\*\*\**^*p <* 0.001. AD/MCI-specific features are framed in blue, and PD-specific features are framed in orange. PD vs. PDM features are framed in red.

The results show that the control group had a significantly greater (*p <* 0.05) average smooth pursuit distance than all other groups, which means that smooth pursuit in this cohort was less interrupted by other eye movements. The AD/MCI and PD groups also exhibited significantly longer (*p <* 0.05) average saccade duration than the control group. These two features were more evident in trial two than in trial one, where the target moved faster. In smooth pursuit trial seven, in which the target pattern covered a larger visual range (infinity sign), the AD/MCI group had significantly larger and more variable (*p <* 0.05) saccade distance, given the mean and standard deviation, compared to CTRL. AD/MCI and PD groups also had significantly higher (*p <* 0.05) velocity than CTRL in trial seven. Overall, participants with AD appeared to have more significant deficiencies in smooth pursuit performance, which seems to support the findings of visual tracking impairments in AD patients by having more interrupted smooth pursuit movements due to catch-up saccades[63].

In Cookie Thief (visual exploration) task, the AD/MCI and PD groups showed significantly higher variability of fixation duration (*p <* 0.05) than CTRL, as shown in Fig. 4. The PD group had a significantly lower (*p <* 0.05) number of fixations than CTRL. Moreover, The PD group differed significantly (*p <* 0.05) from other groups in the standard deviation of saccade duration, indicating a PD-specific feature. Lastly, the relative change in pupil size was shown to be a PDM-specific feature, which was significantly smaller (*p <* 0.05) for the PDM group than other groups in both Cookie Thief and Rainbow Passage tasks. In work [64], pupillary size served as a measure of different attentional demands in local vs. global patterns. Presumably, cognitively demanding tasks (CookieThief, ReadRainbowPassage) require more sustained attention than simple reflexive saccades or antisaccades.

In the reading-out-loud trial of Rainbow Passage, there were no significant differences between people with AD and other groups regarding fixation count, saccade count, and average fixation duration. However, in the silent version, the AD/MCI group had significantly more (*p <* 0.05) fixations, saccades, and a longer average fixation duration than CTRL in Fig. 4. The fixation count was an AD/MCI-specific feature, which also helped differentiate AD/MCI from the PD group. We observed that the rate of fixations and saccades were not significantly different for AD/MCI with other groups because they took longer to finish reading. It is possible that reading and listening to themselves helped the participants with AD visually scan and understand information from the passage in the reading-out-loud trial. The standard deviation of fixation duration was also significantly larger (*p <* 0.05) for NDs groups compared to the control group in both trials of Rainbow Passage.

### B. Task-specific Features

In Fig. 5, we can observe that the PD group had a significantly larger (*p <* 0.05) average difference between the gaze and target positions than the CTRL group in Smooth Pursuit trial two and four, in which targets were moving at a faster speed horizontally and vertically in straight lines. In contrast, no statistically significant differences were observed in trials one and three with a slower target moving speed. In Smooth Pursuit trial five (infinity sign with greater velocity in y direction), the average pursuit gain was significantly greater (*p <* 0.05) for AD/MCI and PDM groups than CTRL, not in the other two infinity sign trials. In Smooth Pursuit trial seven (larger infinity sign), the AD/MCI group carried a significantly larger (*p <* 0.05) standard deviation of the difference between the gaze and target positions than CTRL. The results suggested that larger amplitudes and greater vertical velocity of the target moving pattern increased the difficulty of the task, which could potentially reveal more deficiencies that may be caused by the NDs.

**Fig. 5.**
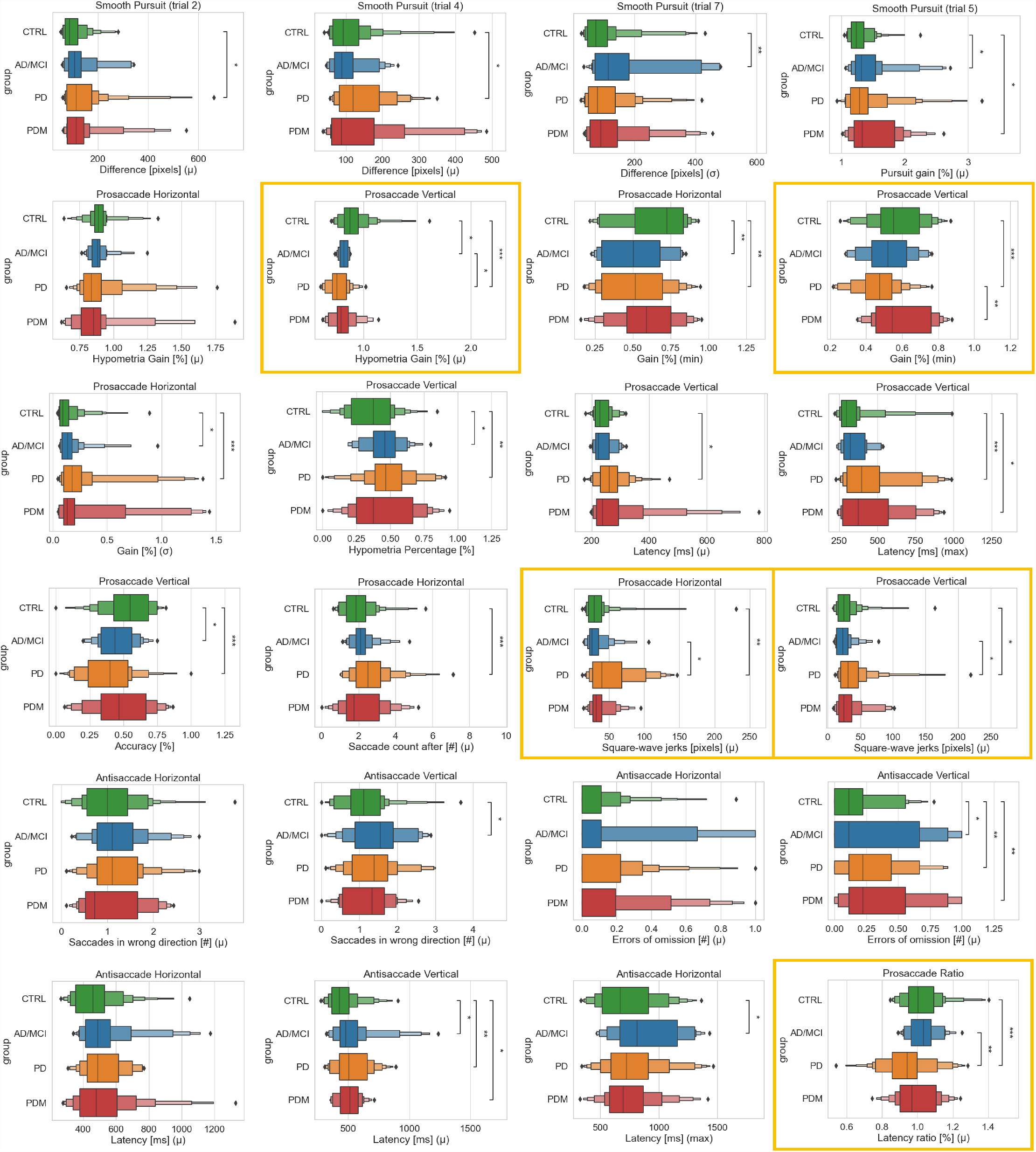
Comparison of task-specific features between CTRL, AD/MCI, PD, and PDM using boxenplots (or letter-value plots [62]). The center line corresponds to the median. The data space is divided further recursively, and more quantiles are added to the depth while the corresponding letter values are reliable estimates. The outliers are represented with a diamond symbol. Asterisks indicate significant differences: ^*\**^*p <* 0.05, ^*\*\**^*p <* 0.01, ^*\*\*\**^*p <* 0.001. PD-specific features are marked with an orange border.

In the horizontal Prosaccade task, Fig. 5 shows that the minimum and standard deviation of gain were significantly different (*p <* 0.05) for the AD/MCI and PD groups than CTRL, while all groups processed a similar average hypometria gain and percentage. Therefore, both groups had a lower magnitude of the shortest hypometria saccade and higher variability of saccade gain. As side-by-side comparisons, both the average hypometria gain and percentage reached the significance level (*p <* 0.05) in the vertical Prosaccade task for AD/MCI and PD groups compared to CTRL in Fig. 5, which means that people with AD/MCI and PD had more trouble reaching the target accurately at their first attempts. The PD group continued to show impaired prosaccade performance regarding the minimum and standard deviation of gain in the vertical direction. These findings could be attributed to a consistent ocular motor abnormality in PD - the saccadic hypometria in which the primary saccade undershoots the target [32]. Also, persons with PD often demonstrate a partially reversible saccadic hypometria which is, in part, dependent upon their L-DOPA state: persons in the optimal L-DOPA ON state may perform differently/better on this task v. L-DOPA OFF state [65], [66]. So the finding here is not static, but rather a dynamic one. Controlling medicine ON and OFF states is one of the future directions.

Similarly, the accuracy for prosaccade and latency in the saccade initiation did not differ significantly between groups for horizontal prosaccades in Fig. 5. However, the PD groups exhibited significantly less accuracy and higher latency in both mean and maximum (*p <* 0.05) for vertical prosaccades than the control group. The accuracy and vertical average hypometria gain are the two most discriminative features, with AUROC of 0.75 and 0.79, respectively, reported in Table IV. The number of corrective saccades after they initiate towards the target until the stimuli disappear is also significantly higher (*p <* 0.05) for the PD group than the control group according to Fig. 5, exhibiting more corrective saccades and less stability in fixation. In addition, the square-wave jerks of the gaze position in the non-dominant direction significantly differed (*p <* 0.05) in CTRL and AD/MCI vs. PD group, which indicates a PD-specific feature. It is possible that people with PD had less control in the less sensible direction while focusing on the main task due to motoric impairments. The latency ratio (horizontal/vertical) significantly differed between PD, AD/MCI, and CTRL, which is also a PD-specific feature. The values unveiled that PD subjects took longer to initiate a saccade vertically than horizontally, while the other groups’ means were centered around 1.0.

For horizontal antisaccades, there was no significant difference in the number of saccades in the wrong direction (prosaccade), between groups. However, for vertical antisaccades, the results showed that the AD/MCI group made more wrong prosaccades relative to the CTRL group (*p <* 0.05) in Fig. 5. Also, the AD/MCI, PD, and PDM groups had significantly more errors of omission, the number of omitted antisaccades relative to a presented stimulus, than the control group (*p <* 0.05) for vertical antisaccades. This scenario means that even if the controls made saccades in the wrong direction, they eventually would make a correct antisaccade, while it did not hold true for some people with NDs, suggesting impairments in detecting an error was made, formulating a correction, and executing it. AD/MCI due to cognitive inattention or bradyphrenia might contribute to responding promptly. A motoric impairment in antisaccade relative to disease severity or L-DOPA state might explain this finding for the PD group. PDM is a mixed cohort with a number of EOM findings which might lead to a reduced detection of positive pathology. We posit that impairments in Cortico-Striatal-Thalamic loop circuits, either subcortically or in the neocortex, may subserve these impairments. There was also a significant difference (*p <* 0.05) in the average latency of the correct antisaccade for all NDs groups compared to CTRL, as shown in Fig. 5. Therefore, the antisaccade task validates itself as a useful tool for measuring the executive function to suppress the reflexive saccade to a peripheral target [31].

### C. Correlation with Clinical Rating Scales

To examine how our extracted features relate to the metrics that clinicians use, we employed the Pearson correlation test [67] to measure the correlation with accepted clinical rating scales, including MoCA and MDS-UPDRS III for AD/MCI and PD subjects. Fig. 6 shows the task-specific features with the most statistically significant correlations with MoCA and MDS-UPDRS III.

**Fig. 6.**
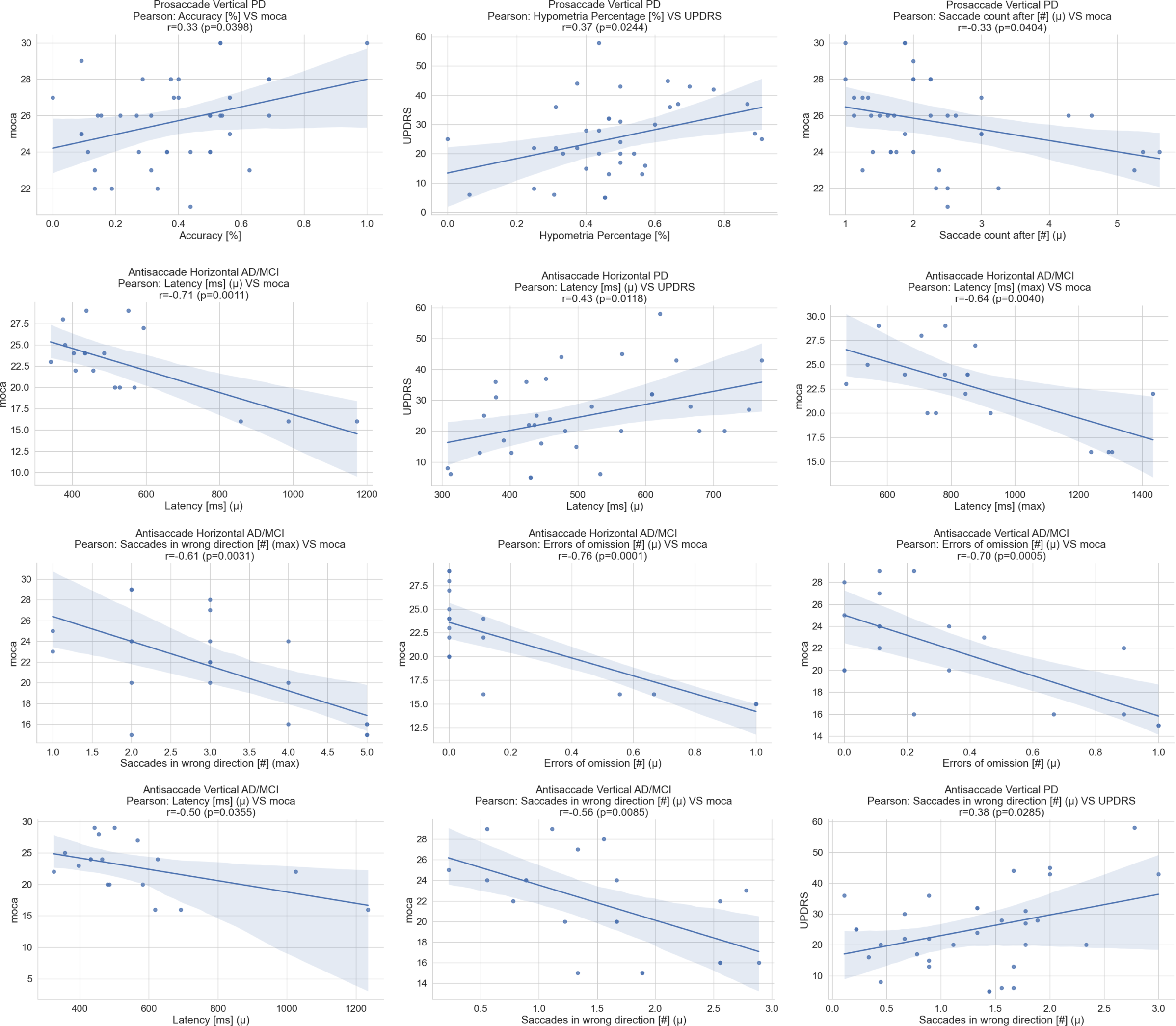
Pearson correlation plots between features and clinical rating scales (MoCA and MDS-UPDRS III). The Pearson correlation coefficient r has values between -1 and 1, which indicates a positive or negative correlation, and essentially is a measurement of normalized covariance between two populations.

Overall, PD subjects with lower MoCA scores tend to have less prosaccade accuracy and more corrective saccades in the vertical Prosaccade task. There is also a positive correlation between hypometria percentage and MDS-UPDRS III for the PD group, which means that PD subjects with higher MDS-UPDRS III tend to generate more hypometria saccades. In the Antisaccade tasks (both directions), AD/MCI patients with lower MoCA scores had longer latency in saccade initiation and more errors due to omissions. From the plots, we can also see that the AD patients had more zero errors horizontally than vertically. The number of saccades in the wrong direction also negatively correlates with the MoCA scores for the AD/MCI subjects. Furthermore, PD patients with higher MDS-UPDRS III scores had longer latency in saccade initiation and more saccades in the wrong direction.

## V. Conclusions and Future work

In this work, we collected eye-tracking data from people with different NDs while performing multiple tasks to assess the motoric and cognitive impairments. A major strength of our multi-modal multi-test rapid battery is portable, and the tasks are fast to complete in contrast to the previously reported versions. We employed signal processing techniques and statistical analysis to design an algorithm for analyzing eye movement and configure a wide range of features that could be used as biomarkers to assist clinicians in accessing NDs. Our algorithm shows promising results in identifying saccade, fixation, smooth pursuit, and blink from eye-tracking data in a single run. Also, besides velocity, accompanying other parameters, such as acceleration, amplitude, and duration thresholds, yields a more comprehensive approach. Moreover, while maintaining interpretability and objectivity, our two sets of features represent valuable metrics to quantify the performance of different visual tasks.

The set of general eye-movement features provides the basic characteristics of eye movement that can be applied to any task. We found that the control group had a significantly greater average smooth pursuit distance than groups with NDs, and the AD/MCI and PD groups were characterized by having longer saccades during smooth pursuit than controls. Also, people with AD/MCI had more variable distances for saccades than controls. These features were more significant, especially when the target was moving faster or covering a larger visual angle. During visual exploration, the variability of saccade duration was much higher in people with PD in comparison to the rest of the groups. The small relative change in pupil size was also unique to the PDM group. While reading a text passage silently, the AD/MCI group made significantly more fixations than the CTRL and PD groups. Therefore, this was an AD/MCI-specific feature. In the reading-out-loud trial, the variability of fixation duration was a PD-specific feature, which could help to differentiate people with PD from controls and people with PD mimicking conditions at the same time.

The task-specific features encode how participants react to the stimuli in specific tasks. Faster target moving speed contributed more significant pursuit difference and the average gain for groups with NDs during smooth pursuit. In the vertical prosaccade task, the PD group showed a significantly smaller average hypometria gain and accuracy, with the most statistical significance and highest AUROC scores of all the features. The minimum saccade gain was also a PD-specific feature and significantly different for PD vs. PDM. Furthermore, people with PD had larger square wave jerks and a smaller ratio of latency (horizontally over vertically) during pro-saccades than AD/MCI and controls. In the vertical antisaccade task, significantly more omitted saccades and longer average latency are reported for the groups with NDs compared to CTRL. The utility of a PDM group was not as an intrinsic/valid construct on its’ own merits but served as the important clinical impact in relation to PD. As the multimodal corpus is an ongoing process, we are expanding our number of participants in each category to balance the experimental groups regarding age, gender, and the number of subjects. By doing so, we expect our findings and generalizability will increase when analyzing more samples in our data set. In the future, this research aims to develop novel machine learning paradigms based on these features to improve differential diagnosis between NDs and monitor disease progression in time. The study of learned features, often non-interpretable, in comparison with these interpretable features, is also a future direction.

## Data Availability

The authors plan to share all data produced in the present study in the future.

## Acknowledgment

This study was funded by the Richman Family Precision Medicine Center of Excellence, the Venture Discovery Fund (VDF), and the Consolidated Anti-Aging Foundation. The study sponsors had no involvement in data collection, analysis, the writing of the manuscript, and submission.

## Appendix

**TABLE III.** The target configurations in different trials in Smooth Pursuit task.

| Trial | X_frequency<br>(Hz) | X_amplitude<br>(°) | Y_frequency<br>(Hz) | Y_amplitude<br>(°) |
| --- | --- | --- | --- | --- |
| 1 | 0.2 | 14 | 0 | 0 |
| 2 | 0.4 | 14 | 0 | 0 |
| 3 | 0 | 0 | 0.2 | 8 |
| 4 | 0 | 0 | 0.4 | 8 |
| 5 | 0.2 | 12 | 0.4 | 6 |
| 6 | 0.4 | 3 | 0.2 | 6 |
| 7 | 0.1 | 18 | 0.2 | 8 |

### Algorithm 1: Eye Motion States Identification Algorithm

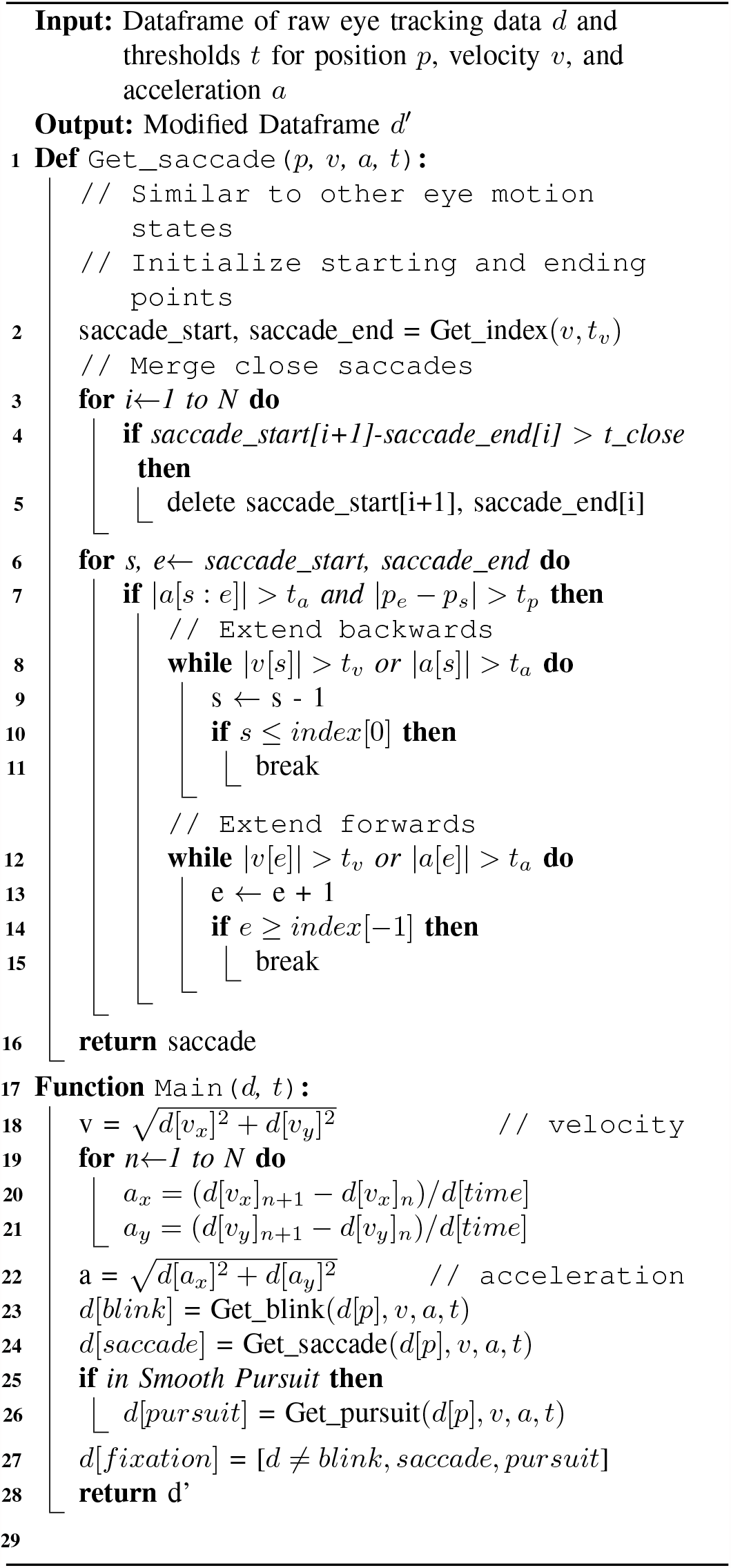

**TABLE IV.**
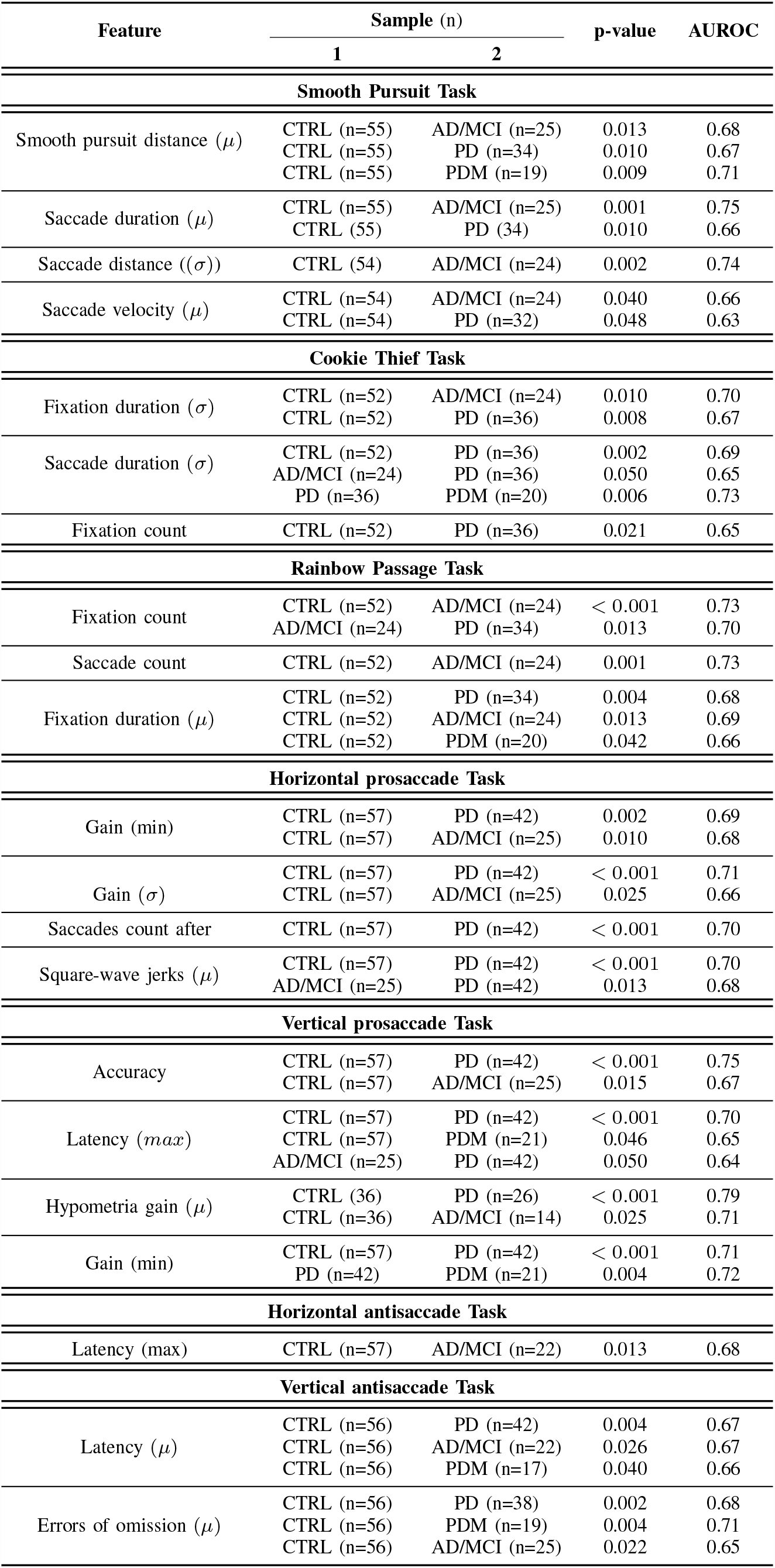
Pairwise Kruskal–Wallis H test results for statistically significant features (*p <* 0.05) from the eye tracking analysis.

## Footnotes

1 The code, including the eye-tracker experiment configuration, is available on [GitHub](https://github.com/Neuro-Logical/Eyetracking_NDs).

